# Descending Brainstem Systems Contribute to Ankle Clonus in Humans with Spinal Cord Injury

**DOI:** 10.64898/2026.05.21.26353256

**Authors:** Etem Curuk, Bing Chen, Alex Benedetto, Matthew Farley, Sina Sangari, Dalia DeSantis, William Z. Rymer, Hans Hultborn, Gregory E.P. Pearcey, Vicki M. Tyselling, Charles J. Heckman, Monica A. Perez

## Abstract

Ankle clonus is a sustained, involuntary, rhythmic muscle contraction frequently observed in humans with spinal cord injury (SCI). Although its pathophysiology remains incompletely understood, converging evidence suggests a role for brainstem systems in its generation. Following SCI, brainstem neuromodulatory inputs partially compensate for the loss of descending motor pathways by regulating motoneuron excitability during involuntary contractions, suggesting their involvement in the generation of clonus. To test this hypothesis, motoneuron excitability in response to Ia synaptic input was quantified using the soleus H-reflex and maximal motor response (H/M ratio), and brainstem involvement was probed using the long-lasting component of the cutaneous reflex (LLR) in the tibialis anterior and soleus muscles, as well as the StartReact response—an involuntary release of a movement triggered by a startling stimulus thought to engage the reticulospinal tract. We studied individuals with chronic SCI, both with and without ankle clonus, using standardized clinical tests across two days. Participants with clonus showed elevated H/M ratios, indicating increased motoneuron excitability, whereas those without clonus exhibited lower values than controls. Additionally, individuals with clonus exhibited longer LLR duration and greater LLR magnitude in both muscles, along with shorter reaction times to startle stimuli, consistent with enhanced monoaminergic and reticulospinal contributions. Notably, LLR duration was positively correlated with both StartReact response and H/M ratio. Together, these findings support a role for descending brainstem systems—particularly monoaminergic and reticulospinal pathways—in the maintenance of clonus in chronic SCI.

## Introduction

Ankle clonus is a common manifestation of upper motor neuron lesions, including those resulting from spinal cord injury (SCI) (Therkildsen et al., 2025). It is characterized by involuntary, rhythmic muscle contractions, typically occurring at 3–8 Hz, most often at distal joints like the ankle (Little et al., 1989; Bravo-Esteban et al., 2013). Clonus can be elicited by passive muscle stretch or during voluntary movement and is associated with reduced quality of life in individuals with SCI (Fleuren et al., 2009; Manella et al., 2013; Khan et al., 2016). Although its underlying mechanisms remain incompletely understood, converging evidence indicates contributions from both central and peripheral systems. This view is supported by findings that clonus frequency can remain stable despite variations in external load (Walsh, 1976; Dimitrijevic et al., 1980) and proprioceptive input (Beres-Jones et al., 2003). In contrast, other studies report that clonus frequency and amplitude scale with externally applied loads (Rack et al., 1984; Rossi et al., 1990), supporting models of reflex instability within a closed-loop sensorimotor feedback system (Hidler and Rymer, 1999, 2000). Despite these insights, the neural mechanisms underlying ankle clonus remain poorly understood.

Motoneuron properties are closely associated with the generation of clonus. Early studies proposed that clonus arises from self-reexcitation within hyperactive stretch reflex pathways (Clare et al., 1951; Cook, 1967; Lippold, 1970; Hagbarth et al., 1975; Nichols et al., 1978; Rack et al., 1984; Latash et al., 1989; Rossi et al., 1990). Computational models have shown that reducing latency in the reflex pathway—either by decreasing conduction time delays or by replacing the soleus slow-twitch muscle characteristics with those of fast-twitch muscles—clonus was attenuated (Hidler and Rymer, 1999). Motoneuron excitability is strongly modulated by monoaminergic neuromodulatory pathways, which facilitate intrinsic depolarizing currents known as persistent inward currents (PICs) (Eken et al., 1989; Heckman et al., 2008; Hultborn et al., 2013). PICs amplify and prolong synaptic inputs, enabling sustained motoneuron firing and prolonged muscle contractions (Hounsgaard et al., 1988; Heckmann et al., 2005). Both animal (Bennett et al., 2001) and human (Gorassini et al., 2004) studies indicate that PIC activation contributes to voluntary and involuntary muscle activity and to the long-lasting components of the cutaneous reflex observed after SCI (Murray et al., 2010; Tysseling et al., 2017). Monoaminergic pathways also enhance the responsiveness of motoneurons and interneurons to fast glutamatergic inputs, such as those originating from reticulospinal pathways (Rekling et al., 2000). Indeed, monoaminergic pathways are thought to amplify postsynaptic potentials generated by reticulospinal pathways in individuals with upper motor neuron lesions, such as those following stroke (McPherson et al., 2018). In individuals with SCI, reticulospinal contributions appear to be further potentiated, particularly in those individuals’ exhibiting spasticity and hyperreflexia (Sangari and Perez, 2020; De Santis and Perez, 2026), underscoring the role of these descending systems in modulating motoneuron excitability after injury. Therefore, we hypothesized that brainstem systems—particularly monoaminergic and reticulospinal pathways—contribute to the regulation of motoneuron excitability and the maintenance of involuntary muscle contractions observed during ankle clonus after SCI.

## Materials & Methods

### Subjects

Thirty-three individuals with chronic SCI (≥1 year post-injury; mean age 47.5 ± 16.0 years; 28 males, 5 females) participated in the study. All participants provided written informed consent, and study procedures were approved by the Northwestern University Institutional Review Board in accordance with the Declaration of Helsinki. Neurological level of injury was determined using the International Standards for Neurological Classification of Spinal Cord Injury (ISNSCI) examination, with all injuries at or above T10. Of the 33 participants, four were classified by the American Spinal Cord Injury Impairment Scale (AIS) as AIS A (motor and sensory complete), one as AIS B (motor complete, sensory incomplete), and 27 as motor and sensory incomplete (AIS C, n = 7; AIS D, n = 20; Table 1). To minimize the effects of antispasmodic or GABAergic medications on muscle tone and neural excitability, participants were asked to withhold these medications for at least 12 h before testing. Clonus was assessed using the drop test and manual stretch test by a trained examiner (Koelman et al., 1993; Manella et al., 2013). The drop test provides reliable measures of clonus that correlate with both the Spinal Cord Assessment Tool for Spastic Reflexes (SCATS) clonus score (Benz et al., 2005) and reflex responses in individuals with SCI (Manella et al., 2013, 2017). Tests were performed in a standardized position (see below) and repeated approximately five times on two separate days. Clonus severity was classified using SCATS criteria (Benz et al., 2005): 0 = no reaction (<1 s), 1 = mild clonus (<3 s), 2 = moderate clonus (3–10 s), and 3 = severe clonus (>10 s). Based on these criteria, participants were classified as Non-clonus (<1 s; 0.35 ± 0.21 s, range 0–0.68 s; n = 14) or Clonus (≥10 s, typically >30 s; n = 19). A clonus burst was defined as an EMG burst followed by a silent period (Figure 1B). The ratio of the soleus maximum H-reflex (H-max) and maximal motor response (M-max) (i.e., H/M ratio) was used as a proxy for motoneuron excitability in response to Ia synaptic input. Brainstem monoaminergic involvement was examined using cutaneous reflexes recorded from the tibialis anterior and soleus muscles, focusing on the long-polysynaptic reflex (LPR) and long-lasting reflex (LLR) components. In addition, the StartReact response—an involuntary release of a planned movement evoked by a startling stimulus and thought to reflect reticulospinal engagement (Tapia et al., 2022)—was assessed in participants with residual voluntary control of the ankle muscles.

**Figure 1.**
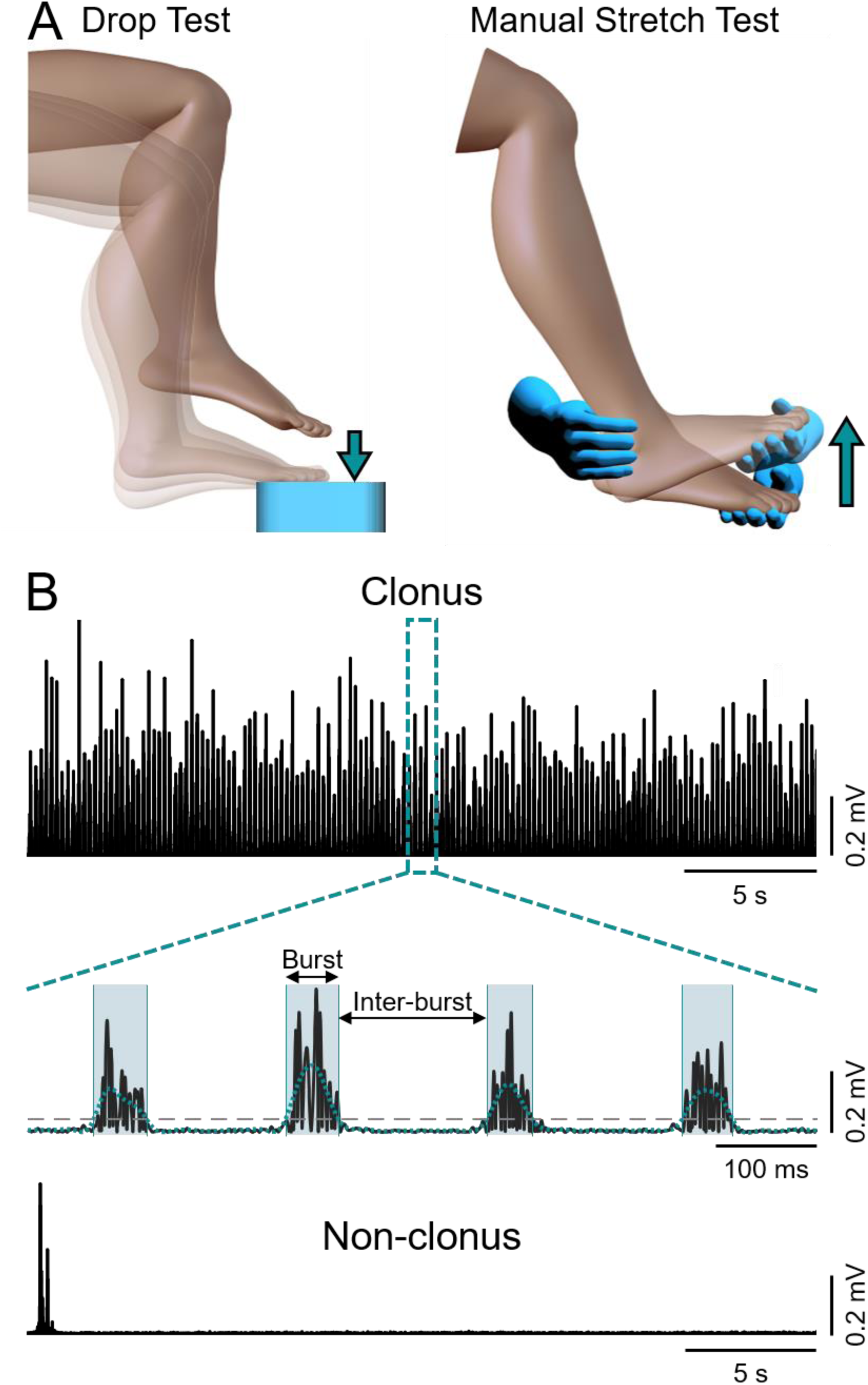
Experimental set-up. **A**, Two clinical tests were used to elicit clonus. The leg drop test (upper left side) involved lifting the leg 10 cm above the platform (blue square) and allowing the edge of the foot to contact the platform. The manual stretch test (upper right side) was performed by the examiner applying a brisk dorsiflexion movement (turquoise arrow). **B**, Rectified electromyographic (EMG) recordings from the soleus muscle in a representative participant with clonus (upper trace), showing rhythmic clonic activity lasting over 30 s. For participants with clonus, clonic EMG activity was quantified by measuring burst duration and inter-burst duration. In contrast, a representative participant without clonus exhibited no rhythmic EMG bursts (lower trace).

**Figure 2.**
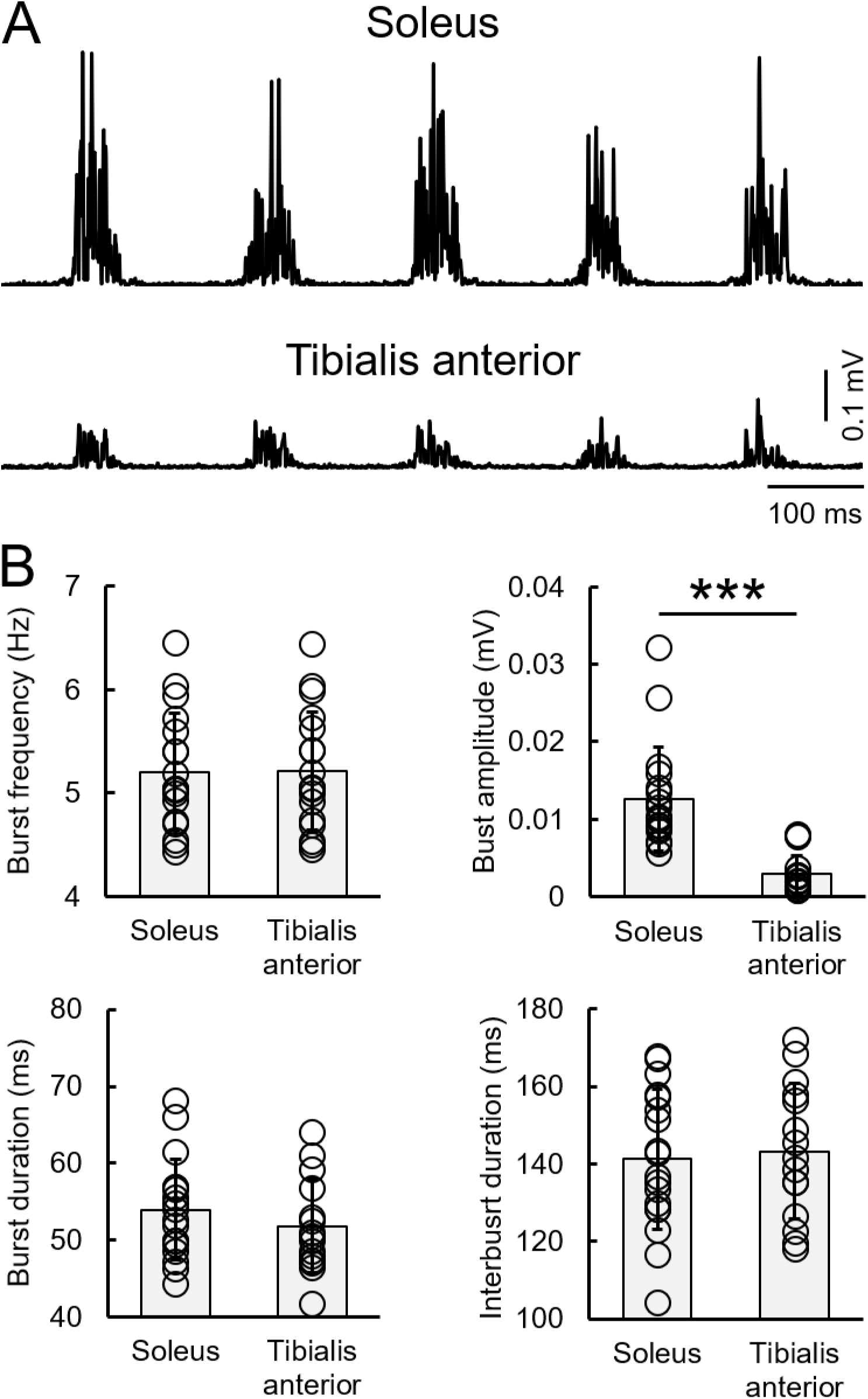
Clonus. **A**, Representative raw EMG recordings demonstrating clonus activity in the soleus and tibialis anterior muscles. **B,** Group data showing in the ordinate burst frequency (upper left), burst amplitude (upper right), burst duration (lower left), and inter-burst duration (lower right) for the soleus and tibialis anterior muscles in participants with clonus. The abscissa indicates the muscle tested (soleus or tibialis anterior). Open circles represent mean values for individual participants for each measurement and muscle. *p < 0.05.

**Table 1.** Spinal cord injury participants. Demographic characteristics of individuals with spinal cord injury (SCI) enrolled in the study. Completion of each study procedure is indicated by an “X.” M, male; F, female; T, traumatic; NT, non-traumatic; AIS, American Spinal Injury Association Impairment Scale; BAC, baclofen; GBP, gabapentin; TIZ, tizanidine.

| Participants | Age (years) | Sex | AIS | Level | Aetiology | Time since injury (years) | Group | Medication(s) | Cutaneous reflex | StartReact | H/M |
| --- | --- | --- | --- | --- | --- | --- | --- | --- | --- | --- | --- |
| 1 | 31-40 | M | C | C8 | T | 10 | Clonus | BAC | X |  | X |
| 2 | 31-40 | M | C | C3 | T | 2 | Clonus | BAC | X | X | X |
| 3 | 51-60 | M | D | C4 | T | 2 | Clonus | BAC | X | X | X |
| 4 | 51-60 | M | D | C2 | NT | 4 | Clonus | BAC,GBP,TIZ | X |  | X |
| 5 | 41-50 | M | D | T9 | T | 4 | Clonus | BAC,GBP | X |  | X |
| 6 | 41-50 | M | B | T8 | T | 8 | Clonus | BAC,GBP | X |  | X |
| 7 | 51-60 | F | C | T10 | NT | 20 | Clonus | BAC,GBP,TIZ | X | X | X |
| 8 | 41-50 | M | D | C7 | T | 14 | Clonus | None | X | X | X |
| 9 | 51-60 | M | C | C5 | T | 12 | Clonus | BAC | X | X | X |
| 10 | 31-40 | M | C | C2 | T | 4 | Clonus | BAC,GAB,TIZ | X |  | X |
| 11 | 31-40 | M | D | T6 | T | 15 | Clonus | BAC | X | X | X |
| 12 | 31-40 | M | D | C5 | T | 22 | Clonus | None | X |  | X |
| 13 | 21-30 | M | D | C6 | T | 3 | Clonus | BAC | X |  | X |
| 14 | 21-30 | M | C | C5 | T | 10 | Clonus | BAC,GBP | X |  | X |
| 15 | 61-70 | F | D | C2 | T | 9 | Clonus | None | X | X | X |
| 16 | 51-60 | M | D | C5 | NT | 22 | Clonus | None | X | X | X |
| 17 | 18-25 | M | D | C4 | T | 4 | Clonus | BAC | X |  | X |
| 18 | 41-50 | F | D | C5 | T | 2 | Clonus | BAC | X | X | X |
| 19 | 51-60 | M | C | C3 | T | 13 | Clonus | BAC, GAB, TIZ |  | X |  |
| 20 | 31-40 | M | A | T4 | T | 17 | No clonus | BAC,GBP | X |  | X |
| 21 | 41-50 | M | A | T5 | T | 7 | No clonus | BAC,GBP | X |  | X |
| 22 | 71-80 | M | D | C5 | T | 5 | No clonus | None | X | X | X |
| 23 | 71-80 | M | D | C3 | NT | 3 | No clonus | None | X | X | X |
| 24 | 41-50 | M | A | T10 | T | 4 | No clonus | BAC,GAB,TIZ | X |  | X |
| 25 | 51-60 | M | D | C3 | NT | 3 | No clonus | BAC | X |  | X |
| 26 | 41-50 | M | D | C5 | T | 4 | No clonus | BAC | X | X | X |
| 27 | 71-80 | M | C | C7 | T | 35 | No clonus | BAC,GBP | X |  | X |
| 28 | 21-30 | F | A | T6 | T | 3 | No clonus | None | X |  | X |
| 29 | 61-70 | M | D | C4 | NT | 15 | No clonus | None | X | X | X |
| 30 | 61-70 | F | D | C1 | T | 8 | No clonus | BAC | X |  | X |
| 31 | 41-50 | M | D | T8 | T | 8 | No clonus | None | X | X | X |
| 32 | 71-80 | M | D | C3 | T | 15 | No clonus | None | X | X | X |
| 33 | 41-50 | M | D | C8 | T | 9 | No clonus | None | X | X | X |

### Electromyographic (EMG) recordings and maximal voluntary contractions (MVCs)

EMG activity was recorded from the tibialis anterior and soleus muscles using bipolar surface electrodes (2-cm diameter Ag–AgCl; ConMed Corp., Utica, NY). Electrodes were placed over the tibialis anterior at one-third of the distance between the fibular head and medial malleolus, and over the soleus on the muscle belly along the midline of the leg. A ground electrode was placed over the patella or medial malleolus. EMG signals were amplified (100 times), band-pass filtered (30–2000 Hz), sampled at 4 kHz, and stored for offline analysis (CED 1401 with Signal software; Cambridge Electronic Design, Cambridge, UK). Participants performed three brief MVCs (3–5 s) in dorsiflexion and plantarflexion, presented in random order and separated by ∼60 s of rest. The maximal mean rectified EMG over a 1-s window was calculated for each MVC, and the average across the three trials was used for analysis.

### Drop test

The drop test is a standardized biomechanical method used to quantify ankle clonus by applying a controlled mechanical perturbation to the plantarflexor muscles (Manella et al., 2013) (Figure 1A). Participants were seated with back support, with the hip and knee flexed to approximately 90°. The metatarsal heads of the test foot were placed on the edge of a 10-cm-high platform, and a padded horizontal bar positioned ∼10 cm above the knee defined the release height. To elicit clonus, the examiner lifted the leg just below the knee until it contacted the bar and then released it, allowing the foot to drop onto the platform and produce a rapid dorsiflexion perturbation. Clonus duration was measured with a stopwatch from the onset of oscillatory movement until cessation or manual termination. Responses lasting longer than 30 s were manually stopped for participant comfort. Each session consisted of five trials, separated by at least 30 s of rest to minimize fatigue.

### Manual stretch test

Clonus was elicited by applying a rapid, manually delivered stretch to the ankle joint (Koelman et al., 1993; Manella et al., 2013). Participants were seated with back support, with the hip and knee flexed to approximately 90°. The examiner held the participant’s foot at the heel while palpating the Achilles tendon and applied a brisk dorsiflexion to the ankle to trigger clonus (Figure 1A).

### Cutaneous reflex

Participants were seated with back support, with the hip flexed to 70° relative to the trunk and the knee flexed to 45° from full extension, and the foot secured with straps using a custom device. The reflex was evoked by electrical stimulation of the medial plantar nerve at the mid-arch using bipolar electrodes (2 cm diameter, 2 cm apart; ConMed Corp., NY). A train of 14 pulses (500 µs, 300 Hz) was delivered via a DS7R stimulator (Digitimer Ltd., UK). We measured the mean amplitude of the LPR (0–500 ms) and LLR (500 ms to reflex offset), as well as LLR duration. Previous studies have reported that LPR and LLR amplitudes in lower-limb muscles plateau at approximately 40 mA for incomplete and 80 mA for complete SCI (DeForest et al., 2020). Consistent with these findings, stimulation intensity was set to 40 mA for the 28 participants with incomplete SCI (18 with clonus, 10 without) and 80 mA for the 5 participants with motor-complete SCI (1 with clonus, 4 without). Reflex onset, offset, and duration were determined from the average EMG across trials. Onset was defined as the point where rectified EMG exceeded baseline by 5 SD over a 100 ms pre-stimulus window and offset as the point where EMG remained below this threshold for at least 500 ms (DeForest et al., 2020). Trials in which reflex offset could not be clearly identified due to sustained single motor unit firing were excluded, as this would affect the LLR duration. The mean amplitude of each reflex component was calculated within a fixed time window corresponding to the longest reflex duration observed across all participants. A total of 20 reflexes were acquired from each participant, and the average EMG was used for analysis.

### StartReact

In the same position described above, the StartReact response was assessed in participants with SCI using a previously established protocol (Baker and Perez, 2017). Participants performed rapid isometric ankle dorsiflexion or plantarflexion in response to a 20 ms light-emitting diode (LED) cue placed ∼1 m in front of them. Visual reaction time (VRT) was defined as the latency from LED onset to rectified EMG onset in the tibialis anterior and soleus. The LED was presented alone or paired with either a non-startling acoustic stimulus (80 dB, 500 Hz, 50 ms) or a startling acoustic stimulus (SAS; 115 dB, 500 Hz, 50 ms) via headphones. Stimuli were delivered in random order with inter-trial intervals of 5–15 s. Visual–auditory reaction time (VART) and visual–startle reaction time (VSRT) were defined as the intervals from stimulus onset to EMG onset. Reaction time was determined as the point where EMG exceeded 5 SD above baseline activity measured over 100 ms pre-stimulus. Each participant completed 20 trials per stimulus condition and contraction type. Three familiarization trials per condition were performed at the start of each session. The VART and VSRT are both mediated via the cochlear nuclei, but only the high-intensity sound of the VSRT activates the reticulospinal pathway (Davis et al., 1982; Brown et al., 1991; Valls-Sole et al., 1999; Tapia et al., 2022). The StartReact effect was estimated as the difference between VART and VSRT. Participants able to perform voluntary contractions participated in the testing (n = 10 clonus; n = 7 non-clonus). StartReact responses were also compared with previously collected data from control participants (n = 19, age 43.3 ± 18.4 yrs; manuscript in preparation).

### H/M ratio

The soleus H-max and M-max were evoked by stimulating the posterior tibial nerve at the popliteal fossa (0.2 Hz) using a constant-current stimulator (DS7R, Digitimer Ltd., UK). Stimulus intensity was increased in 0.05 mA steps from below H-reflex threshold until H-max was reached, then further increased to elicit M-max. M-max amplitude was confirmed at 120% of threshold to ensure full recruitment. The H/M ratio was calculated by dividing H-max by M-max. Values were obtained from a subgroup of participants (n = 18 clonus; n = 14 non-clonus) and compared with previously collected control data (n = 26, age 43.8 ± 14.9 yrs; Chen and Perez, 2022).

### Clonus analysis

Clonus was analyzed in the soleus and tibialis anterior using a custom MATLAB script (v.2020a, MathWorks Inc.) by measuring burst duration, inter-burst interval, mean EMG amplitude, and clonus frequency. In the clonus group, 10 s of EMG activity were analyzed, and trials contaminated by spasm (defined as continuous EMG activity for over 1 s) were excluded. Raw EMG was filtered (20–200 Hz, 4th order IIR), rectified, and smoothed with a 30 Hz low-pass filter. Bursts were identified as peaks in the EMG envelope, with peaks closer than 120 ms (8.5 Hz) excluded based on typical clonus frequencies (4.5–8.4 Hz) (Mummidisetty et al., 2012; Wallace et al., 2012). Burst onset and offset were defined as the timepoint at which the envelope exceeded and fell below the mean threshold for ≥25 ms, respectively. Inter-burst duration was measured from the offset of one burst to the onset of the next, and burst amplitude was calculated as the mean rectified EMG over 20 ms centered on the peak. Clonus frequency was estimated using autocorrelation of the smoothed envelope, defined as the reciprocal of the time lag of the first autocorrelation peak exceeding 0.4. All automated measurements were verified by visual inspection.

### Statistical analysis

Normality of distributions was tested with the Shapiro-Wilk test, and homogeneity of variances with Levene’s test. Sphericity was assessed using Mauchly’s test, and the Greenhouse-Geisser correction was applied when sphericity was violated. Data were log-transformed as needed to meet normality assumptions. Intra-class correlation coefficients (ICC) with 95% confidence intervals were calculated using a two-way mixed-effects model to assess absolute agreement between the drop test and manual stretch test across two days. A split-plot repeated-measures ANOVA was used to examine the effects of MUSCLE (within-subject: tibialis anterior, soleus) and GROUP (between-subject: clonus, non-clonus) on LPR and LLR amplitude, LLR duration, MVCs, and StartReact responses. Within the clonus group, repeated-measures ANOVA assessed the effect of MUSCLE on burst duration, inter-burst interval, burst amplitude, and burst frequency. One-way ANOVA tested GROUP effects on H-max, M-max, and H/M ratio. Pearson correlations were used to examine relationships between LLR, StartReact responses, and H/M ratio. Bonferroni adjustments were applied for post hoc comparisons. Additional analyses compared StartReact responses, H-max, M-max, and H/M ratio in participants with SCI to previously collected age-matched control data. Statistical analyses were performed in SPSS (v26.0; IBM Corp., Armonk, NY), with significance set at p < 0.05. Effect sizes for ANOVA are reported as partial η², representing the proportion of variance in the dependent variable explained by a specific factor, accounting for other variables and error variance. Group data are presented as mean ± SD.

## Results

### Clonus

Figure 1B illustrates rectified EMG traces from soleus muscle. The upper trace shows clonus contractions in a representative participant from the clonus group, lasting over 30 s (only - one portion is shown), while the lower trace shows EMG activity from a participant in the non-clonus group, demonstrating the absence of EMG bursts. Table 2 provides detailed information on EMG burst duration in individual trials for both groups, assessed using the drop test and the manual stretch test, with timing obtained from EMG recordings or visual detection. Most participants in the clonus group (17 of 19) exhibited bursts lasting more than 10 s in all trials, while 2 participants (2 of 19) showed bursts lasting more than 10 s in 90% of trials. In contrast, all participants in the non-clonus group (14 of 14) showed no EMG bursts or clonus activity recorded by a timer in any trial. Reliability testing demonstrated excellent consistency for both assessments: the manual stretch test showed an ICC of 1.0 (95% CI [1.0, 1.0]) between Day 1 and Day 2, and the drop test showed an ICC of 0.97 (95% CI [0.957, 0.98]), suggesting that both assessments provide reliable tests for the presence of clonus.

**Table 2.**
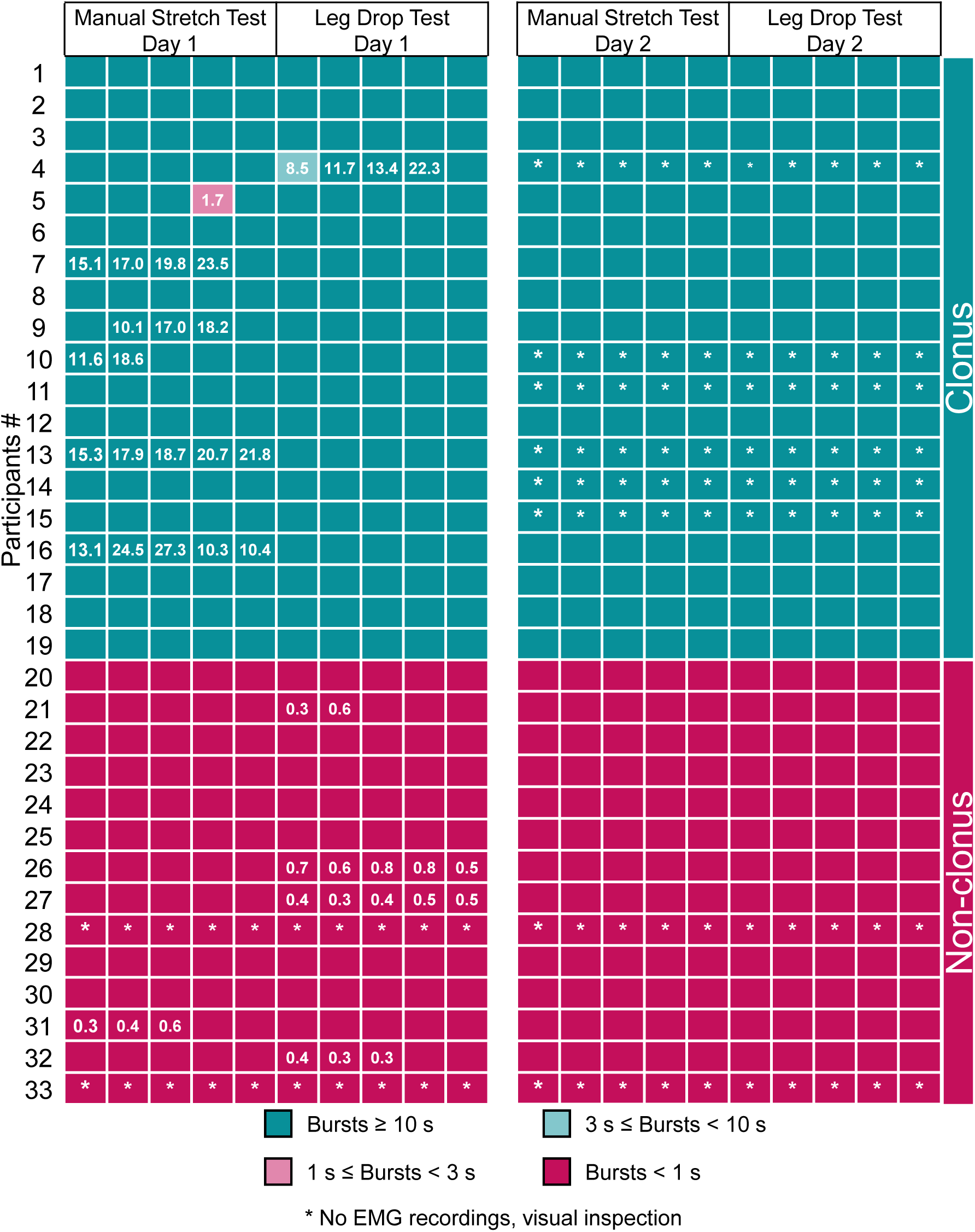
Clonus and non-clonus. Responses obtained during individual trials in participants with clonus (teal) and without clonus (red). Participants were categorized according to the Spinal Cord Assessment Tool for Spastic Reflexes (SCATS) clonus score (Benz et al., 2005). A rating of 0 indicated no reaction (lasting < 1 s; red), a rating of 1 indicated mild clonus lasting < 3 s (pink), a rating of 2 denoted moderate clonus persisting for 3–10 s (light teal), and a rating of 3 represented severe clonus lasting > 10 s (teal). Based on these criteria, participants were divided into two groups: those with average oscillation durations of < 1 s (0.35 ± 0.21 s, range 0–0.68 s; n = 14) and those with oscillations lasting ≥ 10 s (classified as “clonus,” with oscillations in most participants exceeding 30 s; n = 19).

Repeated measures ANOVA in the clonus group revealed a significant effect of MUSCLE on burst amplitude (F₁,₁₄ = 45.16, p < 0.001, partial η² = 0.76), indicating that EMG burst amplitude differed substantially between the soleus and tibialis anterior muscles. Burst amplitude was 0.013 ± 0.007 mV (range 0.006–0.032 mV) in the soleus and 0.003 ± 0.002 mV (range 0.0008–0.008 mV) in the tibialis anterior (p < 0.001). In contrast, MUSCLE had no significant effect on burst frequency (F₁,₁₄ = 0.97, p = 0.34, partial η² = 0.065), burst duration (F₁,₁₄ = 1.12, p = 0.30, partial η² = 0.07), or inter-burst interval (F₁,₁₄ = 1.23, p = 0.29, partial η² = 0.081), suggesting that these parameters were similar across muscles. Inter-burst duration was 141 ± 18 ms (range 104–168 ms) in the soleus and 143 ± 17 ms (range 118–172 ms) in the tibialis anterior (p = 0.34). Clonus frequency was also comparable, at 5.2 ± 0.6 Hz in both the soleus (range 4.4–6.5 Hz) and tibialis anterior (range 4.5–6.4 Hz; p = 0.29).

### Cutaneous reflexes

Figures 3A and 3B illustrate average EMG traces of cutaneous reflexes in the tibialis anterior and soleus muscles from representative participants in the clonus and non-clonus groups. The LLR duration was clearly prolonged in both muscles in the participant with clonus compared to the participant without clonus.

**Figure 3.**
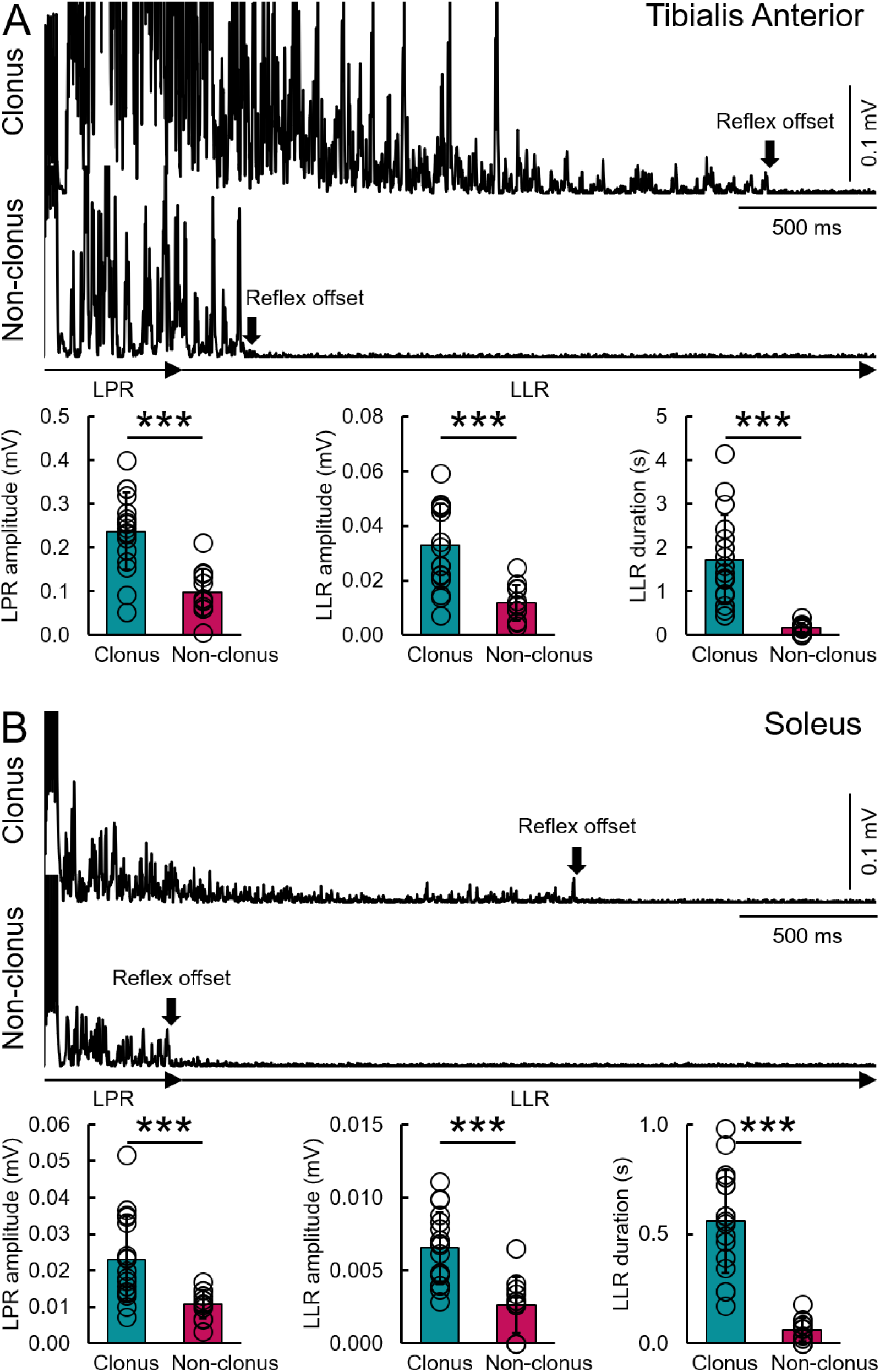
Cutaneous reflex. Raw averaged EMG traces illustrate the cutaneous reflex in an individual with spinal cord injury (SCI) with and without clonus in the tibialis anterior (upper trace, A) and soleus (upper trace, B) muscles. Traces show the long-polysynaptic reflex (LPR) and long-lasting reflex (LLR) components of the cutaneous reflex in both muscles. The duration of the LLR component was calculated starting from 500 ms to reflex offset. Arrows indicate the time of reflex offset for each muscle. Note that LLR duration is shorter in the individual without clonus compared to the individual with clonus for both muscles. Group data are shown for LPR amplitude, LLR amplitude, and LLR duration in the tibialis anterior (A) and soleus (B) muscles. The abscissa indicates the groups tested (clonus, teal; non-clonus, red). Open circles represent mean values for individual participants for each measurement and muscle. *p < 0.05.

We first compared LLR duration and amplitude across groups and muscles. Repeated measures ANOVA revealed significant effects of MUSCLE (F₁,₂₇ = 19.43, p < 0.001, partial η² = 0.42), GROUP (F₁,₂₇ = 40.70, p < 0.001, partial η² = 0.60), and their interaction (F₁,₂₇ = 12.76, p = 0.001, partial η² = 0.32) on LLR duration. *Post-hoc* analyses showed that LLR duration was significantly longer in the clonus group than in the non-clonus group for both the tibialis anterior (1.72 ± 1.02 s vs. 0.17 ± 0.11 s; p < 0.001) and the soleus (0.56 ± 0.24 s vs. 0.06 ± 0.05 s; p < 0.001). Similarly, we found that LLR mean amplitude was significantly influenced by MUSCLE (F₁,₂₇ = 62.39, p < 0.001, partial η² = 0.70), GROUP (F₁,₂₇ = 24.26, p < 0.001, partial η² = 0.47), and their interaction (F₁,₂₇ = 14.15, p = 0.001, partial η² = 0.34). *Post-hoc* analyses showed that LLR amplitude was significantly larger in the clonus group than in the non-clonus group for both the tibialis anterior (32.8 ± 15.3 µV vs. 11.93 ± 6.4 µV, p < 0.001) and the soleus (6.56 ± 2.47 µV vs. 2.63 ± 1.91 µV, p < 0.001). Additional analysis was conducted on LLR mean amplitude normalized to the M-max, it was also significantly influenced by MUSCLE (F₁,₂₇ = 32.08, p < 0.001, partial η² = 0.56), GROUP (F₁,₂₇ = 7.84, p =0.01, partial η² = 0.24), and their interaction (F₁,₂₇ = 7.52, p = 0.011, partial η² = 0.23). *Post-hoc* analyses showed that normalized LLR amplitude was significantly larger in the clonus group than in the non-clonus group for both the tibialis anterior (0.66 ± 0.50 %M-max vs. 0.25 ± 0.10 %M-max, p = 0.007) and the soleus (0.09 ± 0.05 %M-max vs. 0.04 ± 0.04 %M-max, p = 0.01). Within each group, both LLR duration (clonus: p < 0.001; non-clonus: p = 0.001) and amplitude (clonus: p < 0.001; non-clonus: p < 0.001) were larger in the tibialis anterior than in the soleus.

We also compared the LPR (the initial part of the cutaneous reflex) between muscles and groups. Repeated measures ANOVA revealed significant effects of MUSCLE (F₁,₂₇ = 131.7, p < 0.001, partial η² = 0.83), GROUP (F₁,₂₇ = 23.62, p < 0.001, partial η² = 0.47), and their interaction (F₁,₂₇ = 23.24, p < 0.001, partial η² = 0.46) on LPR mean amplitude. *Post-hoc* analyses showed that LPR amplitude was significantly larger in participants with clonus than in those without, for both the tibialis anterior (237.43 ± 88.1 µV vs. 98.36 ± 53.65 µV, p < 0.001) and the soleus (23.02 ± 11.69 µV vs. 10.79 ± 3.83 µV, p = 0.003). Within each group, LPR amplitude was also larger in the tibialis anterior than in the soleus (clonus: p < 0.001; non-clonus: p < 0.001). Additional analysis was conducted on LPR mean amplitude normalized to the M-max, it was also significantly influenced by MUSCLE (F₁,₂₇ = 96.05, p < 0.001, partial η² = 0.79), GROUP (F₁,₂₇ = 11.23, p =0.003, partial η² = 0.31), and their interaction (F₁,₂₇ = 9.86, p = 0.004, partial η² = 0.28). *Post-hoc* analyses showed that normalized LPR amplitude was significantly larger in the clonus group than in the non-clonus group for both the tibialis anterior (4.31 ± 1.79 %M-max vs. 2.23 ± 1.45 %M-max, p = 0.003) and the soleus (0.29 ± 0.18 %M-max vs. 0.15 ± 0.10 %M-max, p = 0.016).

### StartReact responses

Figure 4A illustrates the average rectified EMG traces in the soleus and tibialis anterior muscles for both groups. In the participant with clonus, the StartReact response, calculated as the difference between VART and VSRT, was larger in both muscles than in participants without clonus, suggesting a greater contribution of the reticulospinal pathway.

**Figure 4.**
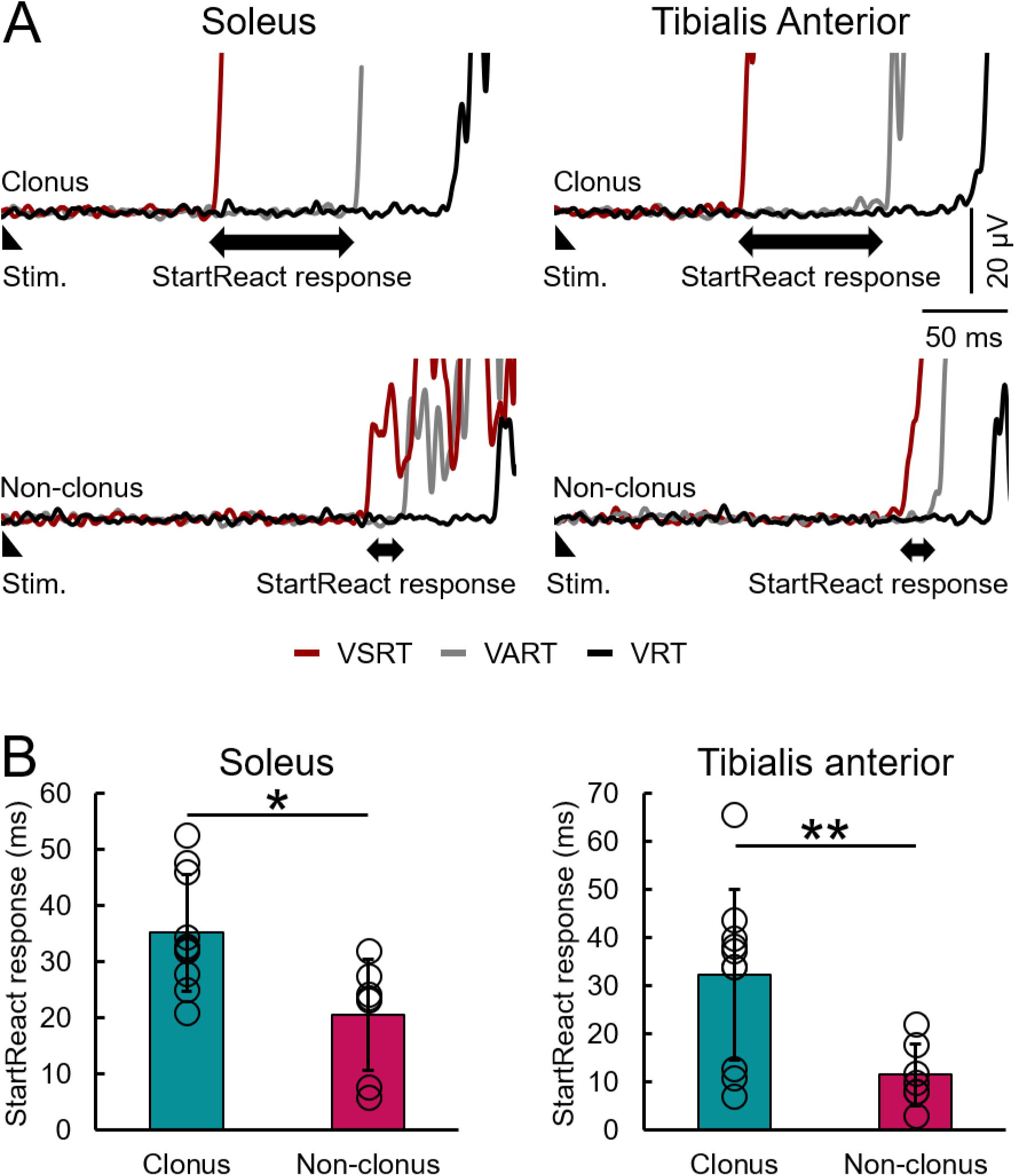
StartReact. **A**, Raw mean EMG traces illustrating visual reaction time (VRT; black), visual–auditory reaction time (VART; gray), and visual–startle reaction time (VSRT; red) in the tibialis anterior and soleus muscles from participants with SCI with and without clonus. For each stimulus condition (VRT, VART, and VSRT) and contraction type (plantarflexion and dorsiflexion), 20 responses were averaged. The StartReact response was calculated as the difference between VART and VSRT (VART − VSRT) in both muscles (double-headed arrow). Notably, in the participant with clonus, reaction time was further reduced and the difference between VART and VSRT was larger in both muscles compared with the participant without clonus. **B**, Group data for the StartReact response in the tibialis anterior and soleus muscles. The abscissa indicates the groups tested (clonus, teal; non-clonus, red). Open circles represent mean values for individual participants for each measurement and muscle. *p < 0.05.

A repeated-measures ANOVA revealed a significant main effect of GROUP on the StartReact response (F₁,₁₅ = 12.21, p = 0.003, partial η² = 0.45), but no main effect of MUSCLE (F₁,₁₅ = 3.00, p = 0.104, partial η² = 0.17) and no interaction (F₁,₁₅ = 0.81, p = 0.38, partial η² = 0.05). *Post hoc* analyses showed that the StartReact response was larger in the clonus group than in the non-clonus group for both the tibialis anterior (clonus: 0.032 ± 0.018 s; non-clonus: 0.012 ± 0.006 s; p = 0.01) and soleus muscles (clonus: 0.035 ± 0.010 s; non-clonus: 0.021 ± 0.010 s; p = 0.02). Within-group comparisons revealed no difference between tibialis anterior and soleus muscles in either the clonus group (p = 1.0) or the non-clonus group (p = 0.10). Comparisons with control participants indicated that the StartReact response was significantly larger in the clonus group relative to controls in both the tibialis anterior (clonus: 0.032 ± 0.018 s; controls: 0.017 ± 0.012 s; p = 0.03) and soleus muscles (clonus: 0.035 ± 0.010 s; controls: 0.022 ± 0.008 s; p = 0.001). In contrast, no differences were observed between the non-clonus group and controls for either the tibialis anterior (p = 0.94) or soleus muscles (p = 1.0).

We also compared the VRT between groups and muscles, which reflects reaction time in the absence of an auditory stimulus. A repeated-measures ANOVA revealed no main effect of MUSCLE (F₁,₁₅ = 2.54, p = 0.13, partial η² = 0.15), no main effect of GROUP (F₁,₁₅ = 0.0004, p = 0.99, partial η² = 0.00003), and no interaction (F₁,₁₅ = 0.03, p = 0.86, partial η² = 0.002) on VRT. *Post hoc* analyses showed that, in the clonus group, VRT did not differ between the tibialis anterior (0.26 ± 0.06 s) and soleus muscles (0.28 ± 0.08 s; p = 0.82). Similarly, in the non-clonus group, VRT was not different between the tibialis anterior (0.26 ± 0.06 s) and soleus muscles (0.28 ± 0.06 s; p = 1.0). Additionally, no differences in VRT were observed between the clonus and non-clonus groups for either the tibialis anterior or soleus muscles (both p = 1.0), suggesting that participants in both groups had a similar ability to response to the LED stimulus in the absence of sound.

### H/M ratios

Figure 5A shows representative raw H-max and M-max traces from the soleus muscle in participants from the clonus and non-clonus groups. A one-way ANOVA revealed a significant effect of GROUP on the H/M ratio (F₁,₃₀ = 49.19, p < 0.001, partial η² = 0.62). *Post hoc* analyses showed that the H/M ratio was higher in the clonus group (0.64 ± 0.12) than in the non-clonus group (0.25 ± 0.18; p < 0.001). Compared with controls (0.49 ± 0.17), the H/M ratio was higher in the clonus group (p = 0.015) and lower in the non-clonus group (p < 0.001).

**Figure 5.**
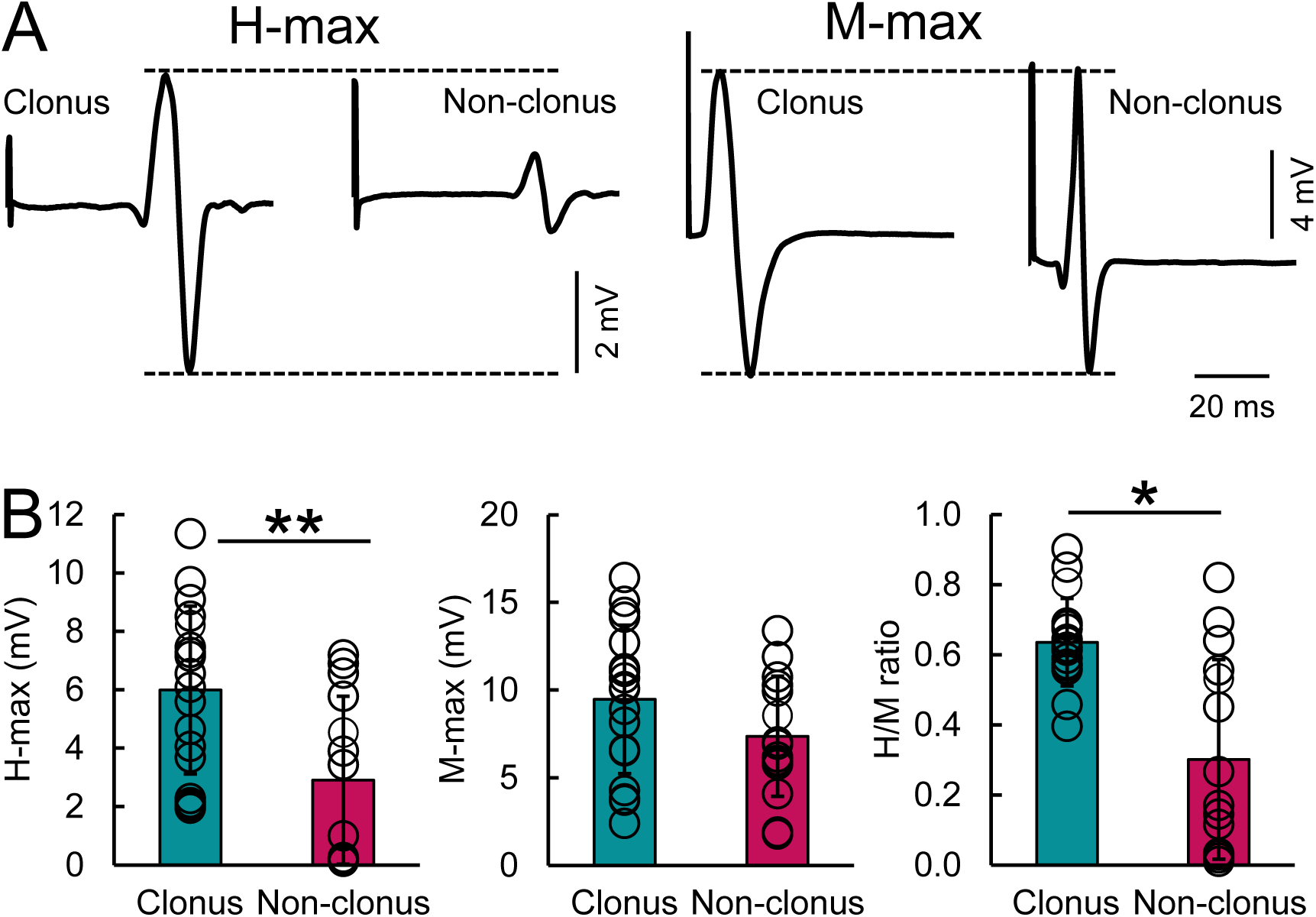
H-max, M-max, H/M ratio. **A**, Representative raw mean EMG traces illustrating the maximal H-reflex (H-max) and maximal motor response (M-max) recorded from the soleus muscle in individuals with SCI with and without clonus. In this example, M-max amplitudes were similar between participants, whereas H-max was smaller in the participant without clonus compared with the participant with clonus. **B,** Group data for H-max, M-max, and the H/M ratio (left to right) in the soleus muscle. The abscissa indicates the groups tested (clonus and non-clonus). The ordinate shows H-max amplitude (millivolts), M-max amplitude (millivolts), and the H/M ratio, respectively. Open circles represent mean values for individual participants for each measurement and muscle. *p < 0.05.

A one-way ANOVA also revealed a significant effect of GROUP on H-max amplitude (F₁,₃₀ = 10.91, p = 0.002, partial η² = 0.27). H-max was greater in the clonus group (5.99 ± 2.87 mV) than in the non-clonus group (2.70 ± 2.67 mV; p = 0.002). Relative to controls (6.41 ± 2.52 mV), H-max was reduced in the non-clonus group (p < 0.001), whereas no difference was observed between the clonus group and controls (p = 1.0). In contrast, no significant effect of GROUP was found for M-max amplitude (F₁,₃₀ = 0.32, p = 0.58, partial η² = 0.01). M-max did not differ between the clonus (9.47 ± 4.24 mV) and non-clonus groups (8.52 ± 5.25 mV; p = 0.58). However, compared with controls (13.23 ± 3.20 mV), M-max was smaller in both the clonus (p = 0.012) and non-clonus groups (p = 0.002).

### Relationship between electrophysiological measures

Figure 6 illustrates the correlations between the StartReact response, LLR duration, and H/M ratio in both muscles. In the soleus muscle, LLR duration was positively correlated with the StartReact response (r = 0.52, p = 0.047; Figure 6A) and with the H/M ratio (r = 0.58, p = 0.001; Figure 6B). In addition, a positive correlation was observed between the StartReact response and the H/M ratio in the soleus muscle (r = 0.69, p = 0.003; Figure 6C). In contrast, no significant correlation was found between LLR duration and the StartReact response in the tibialis anterior muscle (r = 0.30, p = 0.28; Figure 6D).

**Figure 6.**
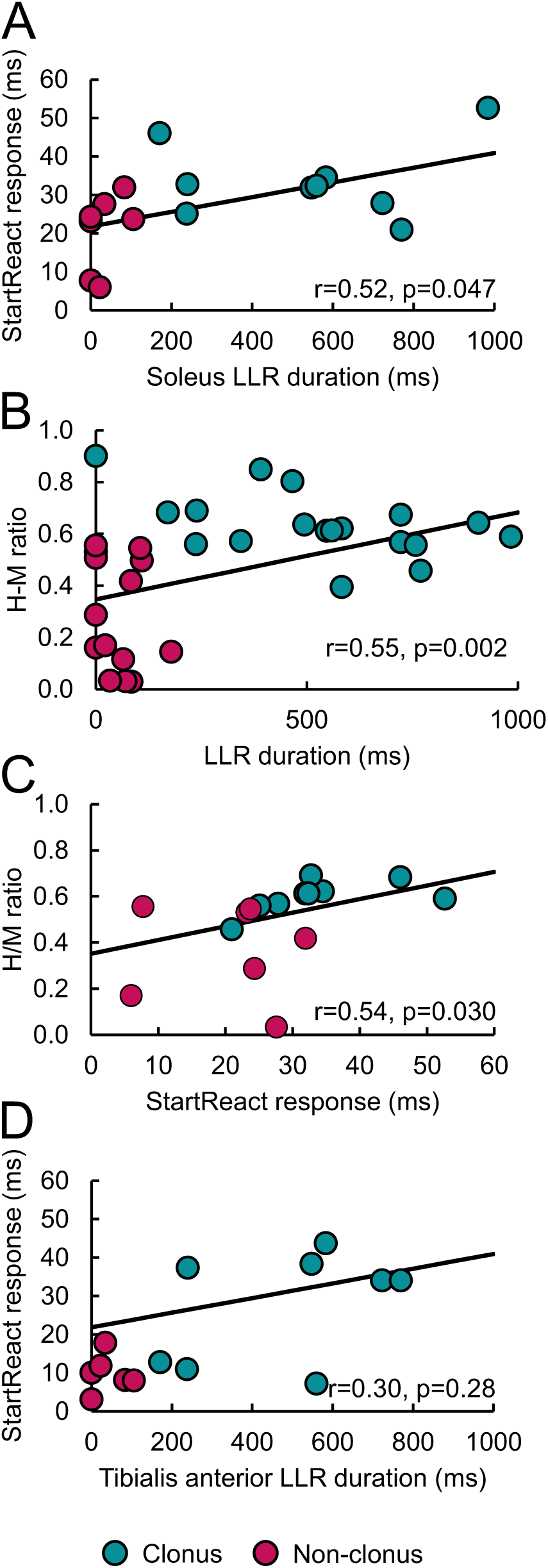
Correlations. Individual data from participants with SCI with clonus (teal) and without clonus (red) are shown. In the upper graphs, the abscissa indicates LLR duration in the soleus muscle, and the ordinate shows the StartReact response (A) and H/M ratio (B). The solid line represents the Pearson or Spearman correlation calculated using all data points from both clonus and non-clonus groups. Notably, LLR duration was positively correlated with both the StartReact response and the H/M ratio. In the lower graphs, the abscissa indicates the StartReact response (C) and LLR duration in the tibialis anterior muscle (D), and the ordinate shows the H/M ratio. *p < 0.05.

## Discussion

Our findings imply that descending brainstem systems play a role in the maintenance of ankle clonus following chronic SCI. Individuals with clonus exhibited elevated H/M ratios, indicating increased motoneuron excitability in response to Ia synaptic input, whereas those without clonus showed smaller H/M ratios than uninjured controls, highlighting the sensitivity of this measure to clonus status. Evidence for descending brainstem involvement was supported by enhanced neuromodulation of the LLR and facilitation of the StartReact response in the clonus versus the non-clonus group. Specifically, participants with clonus demonstrated larger and more prolonged LLRs in the tibialis anterior and soleus muscles, as well as shorter StartReact response latencies, compared with participants without clonus—findings consistent with enhanced monoaminergic and reticulospinal contributions. Notably, LLR duration was positively correlated with both StartReact response and H/M ratio. Together, these findings support the hypothesis that differences in descending brainstem system contributions to motoneuron excitability may, at least in part, explain the presence or absence of ankle clonus following chronic SCI.

### Descending brainstem systems contribute to motoneuron excitability and clonus

Earlier studies have proposed that clonus arises from the self-re-excitation of hyperactive stretch reflex pathways (Clare et al., 1951; Cook, 1967; Lippold, 1970; Hagbarth et al., 1975; Nichols et al., 1978; Rack et al., 1984; Rossi et al., 1990). The co-activation of EMG bursts in soleus and tibialis anterior observed in our study and others (Beres-Jones et al., 2003) is consistent with the emergence of reciprocal facilitation following SCI (Crone et al., 2003), which has also recently been demonstrated in rodent preparations (Rotterman et al., 2024). In line with these observations, H/M ratios are elevated in individuals with clonus compared to both non-clonus participants and uninjured controls (Sangari et al., 2019; Chen and Perez, 2022; Nito et al., 2025). Notably, we also found that H/M ratios are reduced in individuals with SCI who do not exhibit clonus relative to both those with clonus and uninjured controls, consistent with previous reports (Koelman et al., 1993; Manella et al., 2013). A critical question, therefore, is why H/M ratios differentiate individuals with and without clonus, yet do not consistently distinguish those with and without enhanced stretch reflexes (Schindler-Ivens and Shields, 2004; Sangari et al., 2019; Chen and Perez, 2022; Nito et al., 2025). The H/M ratio—defined as the maximal peak-to-peak H-reflex amplitude (a largely monosynaptic spinal response) normalized to the maximal M-wave amplitude (the direct muscle response)—is widely used as an index of motoneuron excitability in response to Ia synaptic input (Pierrot-Deseilligny and Burke, 2005). The stretch reflex, elicited by rapid muscle lengthening (Liddell and Sherrington, 1924), is also commonly used to assess motoneuron excitability in response to Ia input and comprises multiple components, including short-latency monosynaptic, polysynaptic, transcortical, and slower sensory contributions (Birnbaum and Ashby, 1982; Matthews, 1984). Unlike the stretch reflex, the H-reflex is evoked by electrical stimulation of Ia afferents, generating a temporally synchronized afferent volley that enhances spatial summation of excitatory postsynaptic potentials (EPSPs) across the motoneuron membrane (Burke et al., 1983). Consequently, stretch reflex amplitude reflects a combination of temporal and spatial summation of EPSPs, whereas the H-reflex predominantly reflects spatial summation. These differences in synaptic activation may help explain why H/M ratios do not consistently differ between individuals with and without heightened stretch reflexes. Regardless of the methodological approach used to assess motoneuron excitability in response to Ia input in humans, we favor the hypothesis that the observed differences in H-max and H/M ratios between clonus and non-clonus groups primarily reflect differential contributions from descending brainstem systems in these two populations.

To further probe brainstem-derived monoaminergic contributions to motoneuron excitability, we assessed the LLR in the tibialis anterior and soleus muscles. The amplitude and duration of the LLR are largely enhanced by PICs, which amplify synaptic inputs and sustain motoneuron firing, thereby producing prolonged muscle contractions (Hounsgaard et al., 1988; Hultborn et al., 2003; Li et al., 2004; Heckmann et al., 2005). Evidence from both animals (Murray et al., 2010; Tysseling et al., 2017) and human studies (Gorassini et al., 2004; Murray et al., 2010) supports a central role for PIC activation in generating involuntary muscle activity and the LLR following SCI. In our cohort, LLRs were significantly prolonged in participants with clonus compared to those without clonus across both muscles. This observation aligns with previous findings showing that individuals with SCI who experience spasms also exhibit prolonged LLRs (DeForest et al., 2020). The mechanisms underlying clonus may therefore overlap with those contributing to spasms, as both phenomena involve motoneuron hyperexcitability and sustained involuntary muscle activity. Indeed, illustrative examples from several participants (Figure 7) demonstrate transitions in which clonus-like activity evolves into sustained spasms. This transition is consistent with the phenomenon of “warm up” of PICs, in which repeated activation results in progressively stronger PIC amplitudes (Bennett et al., 1998; Gorassini et al., 2002). Another possible mechanism is that movements triggered by long-lasting reflex activity repeatedly reactivate low-threshold afferents, generating additional polysynaptic EPSPs. Through temporal summation and wind-up potentiation, these EPSPs may progressively increase in amplitude, further sustaining rhythmic motoneuron activation. This repeated re-excitation could give rise to clonus, characterized by oscillatory muscle contractions at approximately 5 Hz, consistent with the ∼200 ms duration of individual EPSPs.

**Figure 7.**
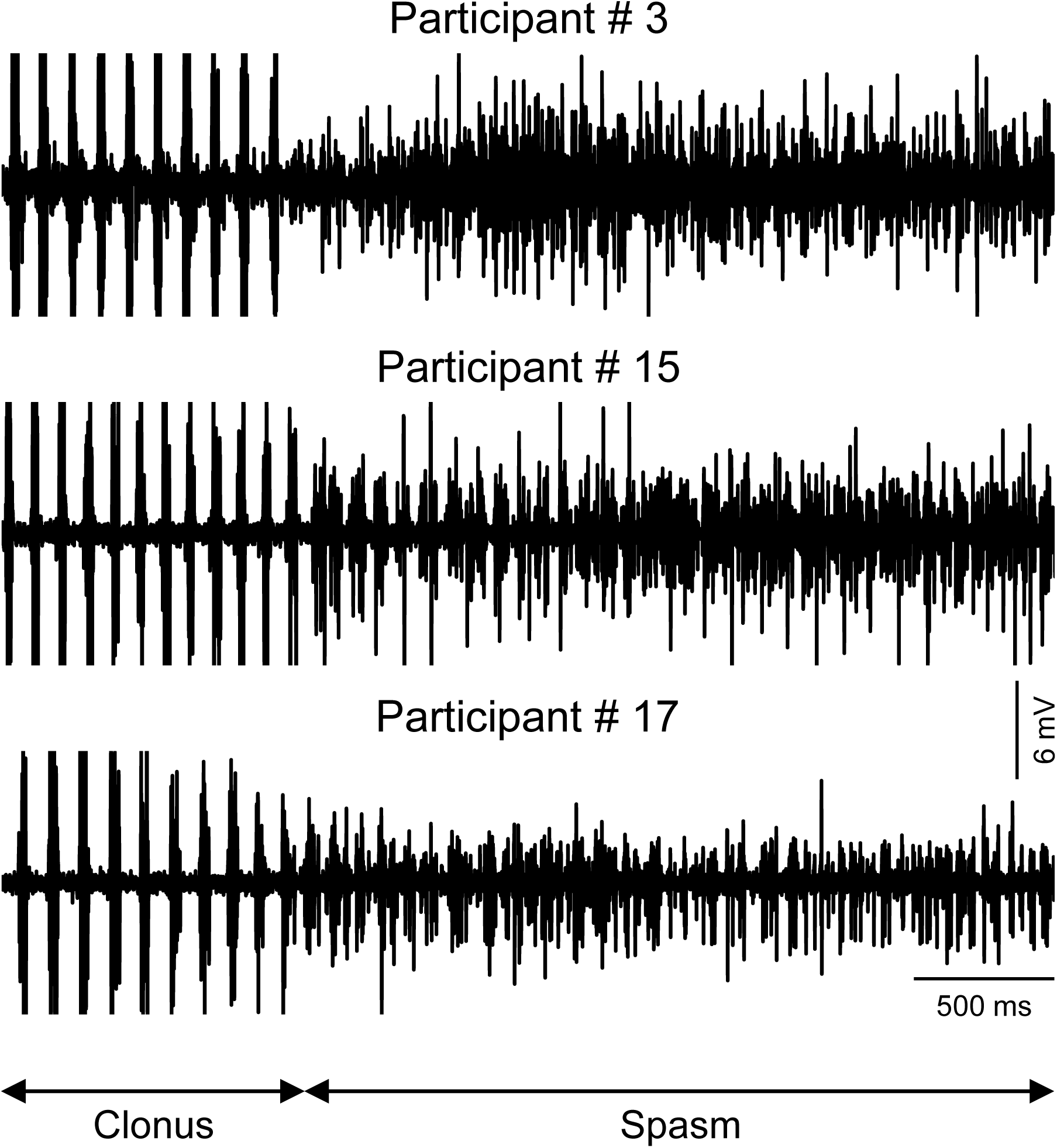
Clonus to spasms. Individual data from participants with SCI and clonus showing representative raw EMG recordings illustrating transitions in which clonus activity in the soleus muscle evolves into a sustained spasm. The representative participants shown correspond to participants 3, 15, and 17 in Table 1. These illustrative examples demonstrate how rhythmic clonus-like activity can progressively transform into prolonged spasmodic muscle activation.

To more specifically probe brainstem contributions via the reticulospinal pathway, we assessed the StartReact response—an involuntary release of a preplanned movement triggered by a startling stimulus that engages the reticulospinal tract (Davis et al., 1982; Brown et al., 1991; Valls-Sole et al., 1999; Tapia et al., 2022). Individuals with clonus exhibited larger difference in reaction times between trials in which the startle stimulus was present compared to trials in which the startle stimulus was not present, consistent with enhanced reticulospinal drive. One possibility is that elevated monoaminergic activity in the clonus group facilitated the StartReact response. This interpretation aligns with evidence demonstrating that serotonin originating from the brainstem enhances the excitability of motoneurons and interneurons, thereby priming them to respond to fast glutamatergic synaptic inputs that drive muscle contractions (Rekling et al., 2000). Consistent with this view, monoaminergic pathways have been shown to amplify postsynaptic potentials generated by reticulospinal motor inputs in individuals with stroke (McPherson et al., 2018). Moreover, reticulospinal contributions appear to be further enhanced in individuals with SCI who exhibit spasticity (Sangari and Perez, 2020; De Santis and Perez, 2026), underscoring the role of descending brainstem systems in modulating motoneuron excitability after injury.

A critical question, however, is why participants without clonus exhibit absent or shorter LLRs. These findings suggest that constitutive activity of serotonin receptors—and the associated activation of PICs—may be reduced or absent in people without clonus. Because both groups exhibited comparable M-max and MVC values, it is unlikely that these differences reflect disparities in overall motor recovery. Experimental in vitro evidence from an incomplete SCI model demonstrates that manipulating brainstem neuromodulatory drive using a selective serotonin reuptake inhibitor modulates LLR activity (Tysseling et al., 2017). Consistent with this, elevated serotonin levels in humans have been associated with clonus (Dunkley et al., 2003). Modeling studies incorporating neuroimaging data further suggest that greater disruption of brainstem-mediated pathways is associated with reduced clinical manifestations of spasticity (Schading-Sassenhausen et al., 2025). Together, these findings support the view that diminished brainstem-derived monoaminergic input may underline the absence or reduction of LLRs in individuals without clonus. However, additional mechanisms cannot be excluded. For example, increased serotonin production within the spinal cord has been demonstrated in animal models of SCI (Wienecke et al., 2014), and serotonin receptor expression on motoneurons increases following SCI (Ren et al., 2013); these changes may differ between individuals with and without clonus. Moreover, inflammatory changes after SCI contribute to altered serotonin receptor activity on motoneurons (Di Narzo et al., 2015), which may also vary between those with and without clonus.

### Functional considerations

Our findings suggest three important clinical implications for individuals with SCI with and without clonus. First, differences in H-max amplitude and H/M ratios between participants indicate that clonus is a sensitive marker of altered motoneuron excitability in response to Ia input. Previous studies have shown that these measures often do not differ between participants with and without other spasticity signs, particularly when defined by increased stretch reflexes (Schindler-Ivens and Shields, 2004; Sangari et al., 2019; Chen and Perez, 2022; Nito et al., 2025). Similarly, measures of Ia afferent transmission regulation—such as post-activation depression at rest (Nito et al., 2025) and presynaptic inhibition during voluntary contraction (Morita et al., 2001; Chen and Perez, 2022) (in communication)—are often comparable in individuals with SCI regardless of spasticity signs. Clonus therefore appears to provide a more objective indicator of motoneuron excitability than commonly used clinical scales, which may lack specificity (Therkildsen et al., 2025).

Second, these findings underscore the importance of differentiating clonus from generalized stretch hyperreflexia when evaluating spasticity in SCI. Although definitions of spasticity vary, increased resistance to rapid muscle stretch is widely recognized as a hallmark feature (Gracies, 2005; Dietz and Sinkjaer, 2007; Nielsen et al., 2007; Dietz and Sinkjaer, 2012). Clinical assessments may be influenced by both neural factors—such as exaggerated stretch reflexes (Lorentzen et al., 2010; Yamaguchi et al., 2018)—and non-neural factors, including increased muscle elastic properties (Mirbagheri et al., 2001) and reduced muscle–tendon unit length (Diong et al., 2012), complicating interpretation (Therkildsen et al., 2025).

Third, standardized assessments like the drop test and manual stretch test show good to excellent test–retest reliability, supporting their use for reproducible clonus quantification (Koelman et al., 1993; Manella et al., 2013). We observed strong agreement between these methods across days but also some variability across trials, suggesting multiple assessments may be needed to accurately determine clonus. Our study included participants with either severe or no clonus, so differences in those with mild or moderate clonus remain to be explored in future studies.

## Data Availability

All data produced in the present study are available upon reasonable request to the authors

